# A one health approach for enhancing the integration of *Salmonella enterica* surveillance in Colombia

**DOI:** 10.1101/2023.03.01.23286234

**Authors:** Johan F. Bernal, Paula L. Díaz, Blanca M. Perez-Sepulveda, María Fernanda Valencia-Guerrero, Magdalena Weisner, Viviana Clavijo, Lucy Angeline Montaño, Stefany A. Arevalo, Ingrid Maribel León, Luis Ricardo Castellanos, Anthony Underwood, Carolina Duarte, Silvia Argimón, Jaime Moreno, David Aanensen, Pilar Donado-Godoy

## Abstract

Foodborne diseases represent a link between environmental, animal, and human health interfaces from the One Health perspective. Whole genome sequencing (WGS) is becoming the gold standard in foodborne surveillance, worldwide. WGS than provides precision data from pathogens allows laboratories to resolve the genetic relations among all sources from One Health perspective, especially during outbreak investigations, getting insights into their transmission routes and pathogenicity. *Salmonella* spp. is the most prevalent foodborne bacteria in Colombia, in 2020, 268 foodborne outbreaks were reported to National health institute (INS) and 3079 invasive and faecal salmonellosis samples from 81% of the Colombian geographical regions. Several studies in different food sources exhibited an increase of contamination with *Salmonella*, a public health concern due to the steady development of antimicrobial resistance associated to specific serovars. However, integration of *Salmonella spp*. data including food-chain supply and clinical interfaces is very scarce in Colombia. This study carried out a national comparison of *Salmonella* isolates collected from food-chain supply and clinical sources. Using an epidemiological and phenotypic approach, we demonstrated the higher resolution of WGS compared with PFGE, routinely used in *Salmonella* surveillance in Colombia. For example, the resolution of PFGE allowed the description of two main clusters of food *Salmonella* Enteritidis isolates which were expanded to eight clades by using WGS. Virulence factors and antimicrobial determinant genes observed in the foodborne clades should be considered a public health concern in Colombia. WGS is a technology that provides precise and valid evidence for the establishment of dissemination routes of foodborne high-risk *Salmonella* clades, but it requires an integrative and continued collaboration between the stakeholders across the One Health interfaces to promote and support integrated real-time actions in public health.

## Introduction

Over 1.7 billion cases of childhood diarrhoeal diseases occur globally every year, causing 525.000 deaths in children under 5 years of age (1). Approximately half of these deaths could be linked to bacterial foodborne illnesses, especially low and middle-income countries (LMICs)(2), where *Salmonella* is one of the four most critical zoonotic bacteria involved in foodborne diseases associated with food production systems (3–6). *Salmonella* occurrence has become a significant global public health concern, due their ability to withstand critical antimicrobials in clinical treatment (7,8).

An effective strategy to tackle the spread of foodborne diseases and antimicrobial resistance (AMR) has been to adopt the One Health (OH) approach of collaboration between members of the food-chain supply, environmental monitoring and the health sector (9,10). Critical levels of AMR in bacteria isolated from humans, the food chain and animal production systems negatively impact the public health and economy at a global level (11–13). There is often a lack of data integration when reports are available for only certain pathogens and others are kept without public access (14). The OH approach recommends establishing a comparable and integrated system to promote more assertive monitoring and measurement strategies (15–17). One strategy for global surveillance of foodborne pathogens and AMR is based on Whole Genome Sequencing (WGS) which is becoming the gold standard method (18)(19). WGS has shown a higher sensitivity, specificity and reproducibility in a shorter time for high-resolution analysis than traditional methods such as pulsed-field gel electrophoresis (PFGE) and ribotyping (20,21). The Global Action Plan (GAP) on AMR in 2015 proposed WGS as a key tool for the integrated analysis of pathogens (19)(22), however most national reference laboratories in Colombia still rely on the use of PFGE for detection of pathogenic bacteria variants.

Colombia proposed the first integrated program in AMR surveillance in Latin-America, the Colombian Integrating Program for AMR Surveillance - COIPARS based on AGROSAVIA. The outcomes from COIPARS showed an estimated prevalence of *Salmonella* in poultry farms (41%) and retail stores (27%), and high levels of AMR to critical antibiotics such as third generation cephalosporins (39%) and fluoroquinolones (29%) (23,24). Similarly, a molecular comparison between poultry farms and retail stores in Colombia revealed the common presence of ESBL/AmpC determinant genes across the interfaces and higher diversity of resistance determinants genes in the food supply chain (25). According to the Colombian National Public Health Institute (INS), *Salmonella* spp. continues to be one of the most important causes of foodborne diseases in the country (26). In 2020, 268 foodborne outbreaks and 3079 salmonellosis cases from 81% (n=30) of all Colombian geographical regions were reported, with 11% of the foodborne illness reports related to poultry products (26). However, the last INS annual report in 2020 showed a reduction of 56% in foodborne outbreaks compared to 2019, this reduction that could be related to the COVID-19 pandemic response in the country (26,27).

A OH approach of surveillance using WGS could generate data relevant to the prevalence, genetic diversity, AMR determinants and mobile genetic elements of pathogens across the food chain and clinical interfaces, making an impact on technology development in health and food safety (25,28–31). This study aimed to provide evidence of the domestic benefits of introducing an integrative genomic analysis (WGS) on *Salmonella* spp. national surveillance. The use of WGS compared to standard PFGE procedure was demonstrated using retrospective data from food isolates from retail stores and clinical isolates from health services between 1997-2017 in Colombia.

## Materials and methods

### Bacterial isolates

The first comparison described as integrated molecular analysis included 801 food and clinical *Salmonella* spp. isolates recovered between 2010-2011. Food isolates (n=118) belonged to the four most prevalent serovars (*S*. Enteritidis, n=61, *S*. Typhimurium, n=14, *S*. Heidelberg, n=20 and *S*. Paratyphi B dT+, n=23) recovered from chicken rinse carcasses at retail stores in 11/32 political divisions (departments) in Colombia were collected between October 2010 to October 2011 by COIPARS-AGROSAVIA (28). Clinical isolates (n= 683) were recovered as part of the National Microbiology Reference Laboratory (NMRL) surveillance program between January 2010 and December 2011 in 28/32 departments in Colombia. These clinical isolates were selected as the same serovars identified for food isolates (*S*. Enteritidis, n=263, *S*. Typhimurium n=263, *S*. Heidelberg n=3 and *S*. Paratyphi B dT+ n=1) and were obtained mainly from blood and stool (29).

The second comparison described as the integrated genomic analysis, the genome of 811 *Salmonella* isolates from food (n=62, COIPARS at AGROSAVIA) and clinical (n=749; NMRL surveillance at INS) were sequenced. The 62 food isolates were selected for sequencing from an initial set of 118 isolates, based on clonal pattern observed using PFGE (DSI=1.0) with clinical isolates. Clinical isolates were selected from the NMRL *Salmonella* surveillance program based on the invasive bloodstream infection outcome report, between 1997-2017. The 151 clinical isolates from all set were selected based on the SNPs analysis defining a probable genetic relation with food isolates. The median fixation rate defined in this study for the core genome in *Salmonella* was three SNPs *per* year, therefore probable related isolates from the clinical surveillance *per* 20 years of surveillance were defined into the ∼60 SNPs distance from a food isolate (32–34).

### Phenotypic characterization

Food isolates were processed in AGROSAVIA following the procedure described by Donado-Godoy et al. (2012). Briefly, an aliquot of chicken carcass rinse was enriched in selective broth, incubated, and the broth plated onto XLT4 agar (Oxoid, UK). Typical *Salmonella* colonies were confirmed at the genus level, and the antimicrobial testing (AST) was performed by BD PhoenixTM automated microbiological system (Becton Dickinson Diagnostic Systems, Sparks, MD, USA). Data interpretation was performed using CLSI breakpoints in 2011 (28).

All clinical *Salmonella* isolates were identified using standard biochemical testing (Triple Sugar Iron Agar (TSI), Citrate, Urea, and motility), and Vitek II system (Biomerieux, USA). Antimicrobial susceptibility testing was performed using the Kirby-Bauer disk diffusion and minimum inhibitory concentrations (MIC) using the MicroScan autoSCAN-4-System (Beckman Coulter) according to the CLSI standards for the corresponding year. All clinical isolates were processed at NMRL at INS. Both food and clinical isolates were serotyped following the Kauffmann-White-Le Minor serological scheme using specific commercial antisera (Difco, United States). Epidemiological and phenotypic data from previous cross-sectional studies and from the National surveillance were integrated to the metadata.

### Molecular and genomic characterization

All 801 isolates were processed as part of PulseNet national surveillance during 2010-2011 in each institution. All isolates were subtyped following PFGE PulseNet-CDC *E*.*coli*-*Shigella*-*Salmonella* standard protocol for *XbaI* enzyme (35). *Salmonella* PFGE patterns were compared using Gel Compare II® (Applied-Maths) software to establish genetic relatedness. Similarity index (SI) was calculated using the DICE coefficient with 1.5% of tolerance in a position of the band of the fingerprinting and the unweighted pair group method by arithmetic mean (UPGMA). *Salmonella* integrated clustering assessment included serovar, PFGE pattern (PP), location and antimicrobial susceptibility profile. The variables, serovars and PP’s were defined as selection measures for the similarity analysis, and binary values were assigned to other variables: (1) when the isolates shared the same output and (0) when the isolates did not share the same output. Pearson Similarity Coefficient (PSC) was calculated using R studio v4.2.0 (36), and a binary similarity score between food and clinical isolates was determined (37–39).

The thermolysates of 811 *Salmonella* isolates were sent to the Earlham Institute (UK) for DNA extraction and sequencing using the LITE pipeline as part of the 10.000 *Salmonella* Genomes consortium, described elsewhere (40). The raw data was transferred to the national participants institutions, and the GHRU bioinformatic pipelines and the computational infrastructure in AGROSAVIA were used for the analysis of the data as OH approach, as described below.

### Bioinformatic analysis

After quality control (QC) using Fastqc (v0.11.8), the raw sequences were trimmed, polished and assembled, and assemblies’ stats were calculated as described in the assembly GHRU-AMR pipeline. Multi locus sequence typing (MLST) and AMR determinants (acquired genes and point mutations) were annotated and summarised in human readable tables following GHRU-AMR pipelines. Plasmids and virulence factors were annotated using ARIBA (v2.14.4) with Plasmidfinder and VFDB databases, respectively. Variant calling was performed using bcftools (v1.9) by mapping to reference genomes (*Salmonella* Enteritidis str. 18569 (NZ_CP011394.1), *Salmonella* Typhimurium str.

LT2 (NC_003197.2) and *Salmonella* Heidelberg str. SL476 (NC_011083.1)), aligned and filtered for low quality base (%QUAL<25 || FORMAT/DP<10 || MAX(FORMAT/ADF)<2 || MAX(FORMAT/ADR)<2 || AX(FORMAT/AD)/SUM(FORMAT/DP)<0.9 || MQ<30 || MQ0F>0.1). Phylogenetic inference was calculated using IQtree (1.6.8) with GTR+G model and 1000 bootstrap replicates. All details of the GHU-AMR pipelines and parameters used in the analysis are described at https://www.protocols.io/view/ghru-genomic-surveillance-of-antimicrobial-resista-bpn6mmhe. SNPs matrix heatmap was performed using R studio (v4.2.0) with the library ComplexHeatmap (41). Clade definition was based on the SNPs matrix generate by the phylogenetic analysis in each serovar using rhierbaps (42) with default settings in R studio (v4.2.2). Epidemiological, phenotypic and genomic data were integrated in a datasheet (.csv) using R studio (v4.2.0). All results were uploaded to the Microreact platform and visualized as a public access dynamic report (43).

### Availability of sequence data

The raw sequencing data generated from this study were deposited at the EMBL European Nucleotide Archive (ENA) repository under the project accession numbers PRJEB35182 and PRJEB47910.

### Ethical considerations

The ethics and methodologies committee for research (CETIN/CEIN) of National health Institute of Colombia waived the ethical approval for this study.

## Results

Evaluation of variables (serovar, PFGE pattern, location, susceptibility profile) showed a correlation between 41% (48 out of 118) food isolates with clinical isolates between 2010-2011. The Pearson similarity coefficient (PSC) for all isolates was defined at 0.87 (p>0.000), stating a strong correlation between food and clinical isolates (Table S1). The phylogenetic inference from the 811 *Salmonella* spp. isolates revealed that all food isolates were genetically related to previously reported clinical isolates from the national surveillance program between 1997-2017. Phylogenetic inference by serovar of 213 closely related food and clinical isolates (*S*. Enteritidis, n= 167; *S*. Typhimurium, n= 35; *S*. Heidelberg, n=11) provided a better resolution of the genetic relationship between interfaces, defining the high-risk foodborne clades in each serovar (Table S2). Of all *Salmonella* spp. isolates compared, 25% clustered into the 12 high-risk foodborne clades described in this study.

### Salmonella Enteritidis

#### Relatedness analysis

*Salmonella* Enteritidis isolates from the food interface (n=61) showed eight unique PFGE patterns (PPs), of which ICA JEG.X01.0001 (n=28) and JEG.X01.0006 (n=27) clustered 90.2% of isolates with a Dice Similarity Index (DSI) >0.96 (figure 1a). Three food PPs were indistinguishable from a unique clinical PPs in the PulseNet national database (DSI=1.00; figure 1b). The two most frequent food (PPs, JEG.X01.0001 - JEG.X01.0006, 55/61; 83%), and clinical PPs (JEG.X01.0001 - JEG.X01.0038, 230/263; 87%) exhibited clonal relatedness (DSI=1.00; figure 1b), defining Cluster-1 (INS JEG.X01.0001 - ICA JEG.X01.0001) and Cluster-2 (INS JEG.X01.0038 - ICA JEG.X01.0006). An additional food singleton PP JEG.X01.0008 was found clonally related to clinical PP JEG.X01.0068 (figure 1b). Cluster-1 *S*. Enteritidis food isolates were recovered from Bogotá D.C (92%), Atlántico (4%) y Arauca (4%). Cluster-1 clinical isolates were collected from several departments (n=14) across the country, especially from Bogotá D.C (28%) and Atlántico (1%), and non-isolate was collected from Arauca. All Cluster-2 food isolates were collected from Bogota, whereas clinical isolates were spread to several departments (n=12), as Bogotá (30%). Integrating the four epidemiological variables (serovar, PFGE pattern, AST and location) from 61 food and 263 clinical *Salmonella* Enteritidis isolates revealed a strong correlation between food and clinical isolates with a PSC of 0.86 (p<0.000). This correlation was explained by the clonal behaviour of 42 food and 54 clinical isolates, all of them pan-susceptible to tested antimicrobials and recovered from Bogotá, Valle del Cauca and Risaralda during 2010-2011 (Table S1).

**Fig 1.**
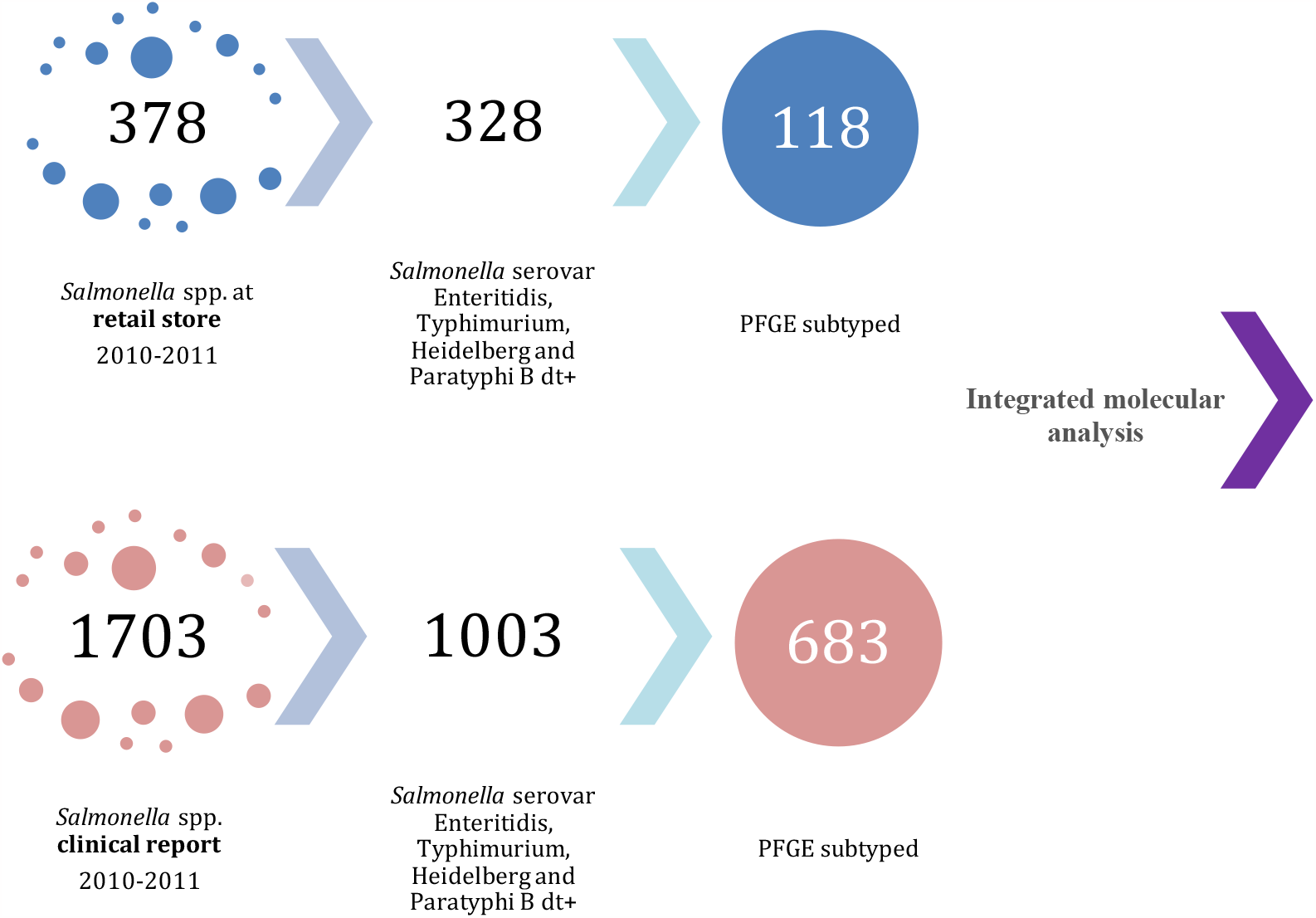
Distribution and selection of samples from food and clinical interfaces included in the molecular integrated analysis

From WGS data, an initial alignment of single nucleotide variants was generated with 394 sequences (food (n=45) and clinical (n=349)) mapped to reference (NZ_CP011394.1). A subset of 167 (42%) genomes were defined as related isolates, including food (n=45) and clinical (n=122) isolates (figure 1c). Sequence types from the subset of *S*. Enteritidis isolates were described into the Complex Group (CG) 11 including ST11(99%) and the novel ST 9293 (0.6%) reported for first time in this study (Table S2). A total of 8 *Salmonella* Enteritidis Clades (SECs) were defined based on SNPs distance and the tree clustering (figure 1c) and visualized on heatmap (figure 1d). Clades analysis by WGS exhibited more than four folds more discriminatory resolution in the same population than the previously described clonal PFGE Cluster-1 and Cluster-2 (Table S2). All SEC1 isolates were recovered from Bogotá between 2010-2012, the majority (8/11) of which were food isolates, including two close zoonotic events (< 2 SNPs) identified during 2010-2011. SEC2 comprised 18 isolates recovered from seven departments between 2010 and 2016. A close genetic event (<2 SNPs) reported in two departments (Bogotá and Risaralda) during 2010 was first observed, after which other clinical isolates were reported for years 2012,2014 and 2016 (Table S2). Two sub-groups of isolates were observed in SEC3 (n=34) differentiated by 13 SNPs of distance, showing food-clinical close genetic events (1-2 SNPs). Clinical isolates were mostly clustered in sub-group SEC3.1 whereas food isolates clustered in SEC3.2, both isolated between 2009-2016. SEC4 isolates (n=9) were collected between 2006-2011 in 4 departments, of which a close genetic event was identified in a multi-department source between 2010-2011. The majority (94%) of SEC5 isolates were from clinical origin, and two food isolates showed relatedness with 6-14 SNPs of distance with clinical isolates between 1998 and 2016. SEC6 was composed of mostly (96.4%) clinical isolates collected from nine departments between 2004 to 2015, with one genetically related food isolate (< 6 SNPs). All SEC7 isolates (n=12) were recovered from clinical between 2002-2016 from 7 departments. SEC7 was resulted a close well-differentiated clade to SEC8 by 28 SNPs of distance with common ancestry. Finally, SEC8 composed of 21 isolates was recovered from 6 departments between 1999 to 2014. A genetically close event of food isolates with a clinical isolate (4 SNPs) was observed during 2010-2012. Overall, SECs isolates were recovered from 19 departments, across Colombia, between 1998-2016 (figure 1c; Table S2).

#### Antimicrobial resistance

All related isolates from the two clusters and the singleton were susceptible to all antibiotics tested (Table S1). *S*. Enteritidis food isolates (65.3%) non-related to clinical isolates were resistant to one or more antibiotics that included β-lactams, quinolones, fluoroquinolones, tetracyclines, sulfamethoxazole/trimethoprim, nitrofurantoin and amoxicillin/clavulanic acid. The Multi-Drug Resistance (MDR) profile AmcAmpCzoCtxFoxCroNaCipNitTe was found in two food isolates non-related to clinical isolates from the department of Arauca and the city of Bogotá. Other less complex AMR profiles AmcAmpSxtTe and AmcAmpCtxNaGen were found in two clinical isolates non-related to food interface from departments of Antioquia and Nariño. Most *S*. Enteritidis isolates (89%) did not carry AMR-determinant genes, and the clinical-food close related isolates were susceptible to all antimicrobials tested. Twelve isolates (11%) from different SECs (1-8) carried AMR determinant genes to aminoglycosides, ß-lactams, trimethoprim, sulphonamides, quinolones and tetracycline. Incompatibility groups IncN_1 and IncI_1 were found related to MDR clinical isolates from SECs 3, 5 and 6 (Table S2).

#### Virulence factors

Virulence factors described previously in the *Salmonella* genus were found in almost all isolates (99%) of S. Enteritidis encoding partially *Salmonella* pathogenicity island SPI-1 and SPI-2, including two Type 3 Secretion Systems (T3SS1-T3SS2) with multiple exclusive and translocate effectors, and one Type 8 Secretion System (T8SS). Additionally, adherence, membrane, flagellar, chemotaxis kinase operons, and metal homeostasis pathways genes were described for all isolates (Table 2). Clinical-food close related events exhibited the same virulence repertoire across all SECs. Other virulence genes related to mobile genetic elements (MGE) such as ybtE, ybtP, ybtU, fyuA, gogB, irp1, irp2 and gtrB were observed in some isolates from SECs 2, 3 and 7. Also, RND efflux system gene acrA and T3SS effector sspH1 related to AMR in *Salmonella* spp. were sporadically observed (Table S2).

All results of *Salmonella* Enteritidis could be visualized at the microreact project (https://microreact.org/project/hjw5geMmKWAAgKxhA9Y3QX-salmonellaentericaenteritidis-from-the-national-surveillance-in-colombia-as-one-health-approach)

### Salmonella Typhimurium

#### Relatedness analysis

*Salmonella* Typhimurium isolates from food (n=14) showed nine unique PPs with DSI values between 0.72-1.0, the most frequent food PPs including JPX.X01.0006, JPX.X01.0008 and JPX.X01.0009 showed DSI values above 0.76 (figure 2a). Additionally, food PPs JPX.X01.0008 and JPX.X01.0007 were found related (DSI = 0.86) (figure 2a). Two unique PPs from food interface JPX.X01.0004 and JPX.X01.0007 (6/14) were indistinguishable from two unique clinical interface PPs JPX.X01.0197 and JPX.X01.0237 at the PulseNet National database (9/416), respectively (DSI=1.0, figure 2b). The PPs JPX.X01.0008 and JPX.X01.0005 from food origin were included as additional cluster in the comparison due the close genetic relation with the clinical clonal related PP JPX.X01.0007.

**Fig 2.**
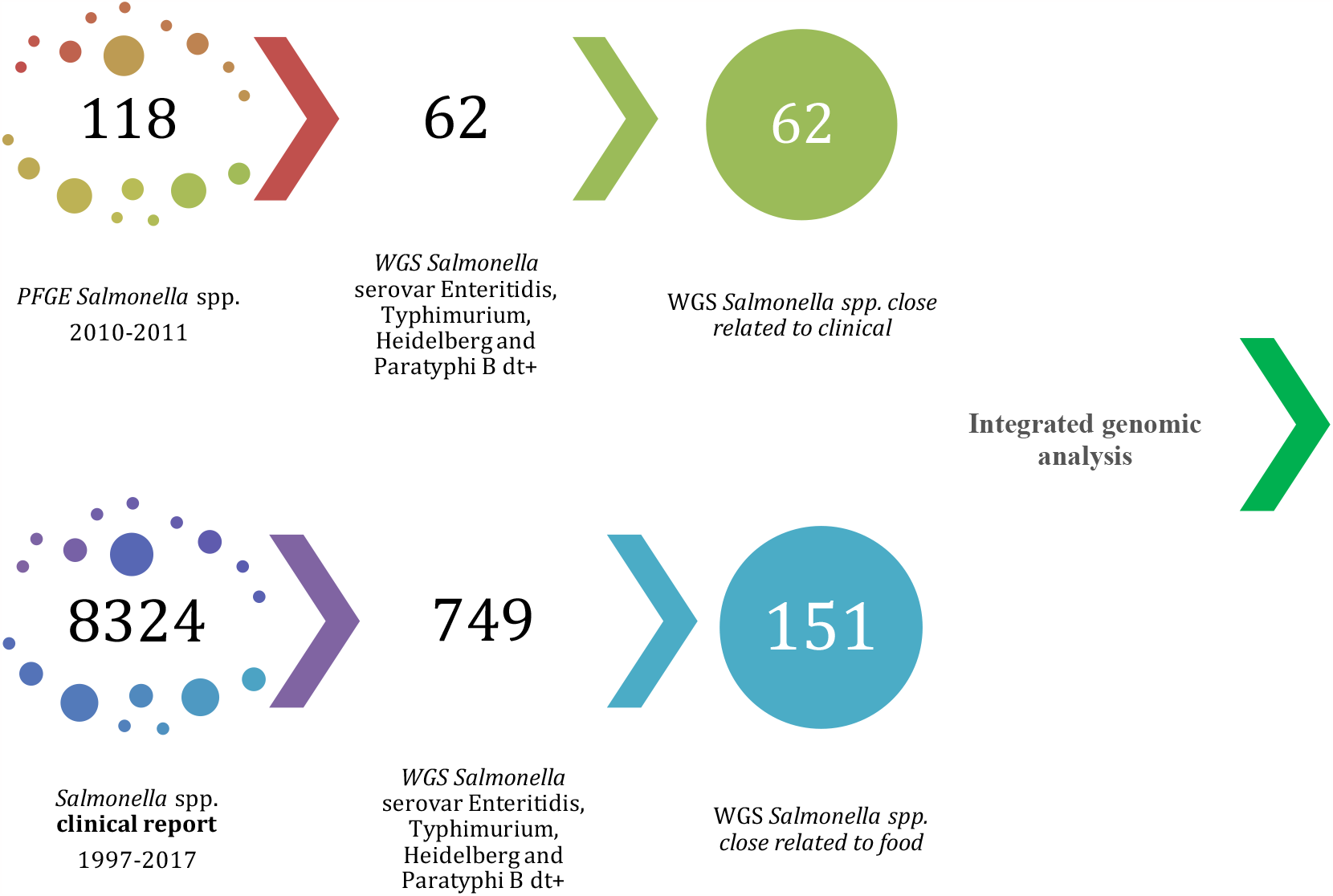
Distribution and selection of samples from food and clinical interfaces included in the genomic integrated analysis

*S*. Typhimurium Cluster – 1 (ICA JPX.X01.0008 – ICA JPX.X01.0005), Cluster -2 (ICA JPX.X01.0007 - INS JPX.X01.0237) and Cluster -3 (ICA JPX.X01.0004 - INS JPX.X01.0197) were defined. *S*. Typhimurium food and clinical isolates belonging to Cluster 1 were recovered in Antioquia (1/2) and Valle del Cauca /1/2), to Cluster-2 were recovered in Bogota (4/4) and to Cluster-3 were recovered in Bogotá D.C (3/12), Antioquia (6/12) y Boyacá (3/12). Other food isolates non-related to clinical interface were recovered from Bogotá (8/14), Antioquia (2/14), Tolima (1/14) and Valle del Cauca (1/14). Four epidemiological variables from 14 food and 416 clinical *S*. Typhimurium isolates were compared, defining PSC in 0.89 (p<0.000). Comparison demonstrated clonal behaviour in 2 food and 4 clinical isolates with AMR profiles CAmcAmpTeStr and TeStr from Bogotá during 2010-2011 (Table S1).

In WGS, the variants calling in *S*. Typhimurium were generated mapping to reference (NC_003197.2) a total of 267 pair sequences: Food (n=5) and clinical (n=262). A group of 35 food-clinical isolates (13%) were selected based on SNPs distance (∼60 SNPs) for the latest phylogeny analysis: Food (n=5) and Clinical (n=30) isolates were recovered between 1999-2016. All sequence types were found belong to Complex Group 19 including ST’s 19 (94%), 7423 (3%) and 7617 (3%), and geographically distributed in 8 Colombian departments (Table S2; figure 2c). From 35 selected isolates, a total of 3 *Salmonella* Typhimurium Clades (STCs) recovered across the country were labelled as probable foodborne (Table S2). Phylogenetic inference described a similar clades distribution to previously observed in the PFGE molecular analysis: STC1 (Cluster -1; ICA JPX.X01.0008), STC2 (Cluster -2; ICA JPX.X01.0007-INS JPX.X01.0237) and STC3 (Cluster-3; ICA JPX.X01.0004 -INS JPX.X01.0197). Nevertheless, other PPs were included into the defined clades. STC1 was the most frequent clade (n=14; 40%) recovered from five departments between 1999-2016 and the majority (11/14) was isolated from clinical. STC1 isolates exhibited no close genetic events, but three related events (< 26 SNPs) were observed during 2003-2016 (figure 2c). STC2 was less frequent clade (n=8; 23%) recovered from four departments between 2004 and 2010; One zoonotic event (7-9 SNPs) was detected in 2004 and 2006. At last, STC3 isolates (n=13; 37%) were recovered from four departments between 2007-2015, a close related event was established with 10 SNPs of distance, during 2007 and 2010 (Table S2).

#### Antimicrobial resistance

All food isolates from Cluster -1, -2 and -3 showed resistance to at least one antibiotic family: ß-lactam, ß-lactamase inhibitor, streptomycin, chloramphenicol and tetracycline (Table S1). Multi-drug resistance profile CAmcAmpTeStr / AmcAmpTeStr were found in four isolates from Bogotá belonging to Cluster-2 (Table S2). Other AMR profiles in Cluster-1 remitted from departments of Antioquia (50%), Boyacá (25%) and Bogotá (25%) were observed: NaTeStr (5/12), TeStr (6/12) or Te (1/12) (Table1). Resistance profile TeStr was observed on a unique Cluster-2 clinical isolate from Bogotá, in contrast, three food isolates from PP JPX.X01.0008 in Huila, were susceptible to all tested antibiotics (Sup. Table 2). Specific blocks of AMR determinants genes were observed in each STCs. All STC1 isolates were multidrug-resistant including several antibiotics families: aminoglycoside, ß-lactam, trimethoprim, sulphonamides, macrolide, quaternary ammonium, phenicols, colistin, quinolones and tetracycline. Three different AMR genetic platforms were observed, standing out the presence of colistin resistance determinant gene mcr-5.1 in food isolates. The presence of plasmid incompatibility groups colpVC, colRNAI and IncC_1 was frequently identified in isolates from STC1. Similarly, STC2 isolates exhibited AMR determinant genes to aminoglycosides, ß-lactams, phenicols, quaternary ammonium, sulphonamides and tetracycline, configured in a common block of AMR genes. All isolates from STC2 carried IncFIIB and IncFII_S plasmids. Finally, STC3 isolates exhibited blocks of AMR determinants to aminoglycoside, sulphonamide and tetracycline but five isolates exhibited additional quinolone resistance. IncQ1 and IncX1 plasmids were identified in all isolates and col156 plasmid was frequently observed. Co-location of qnrB gene and Col440I incompatibility group was defined conferring resistance to quinolones in *Salmonella* foodborne isolates from food and clinical interfaces (Table S2).

#### Virulence factors

Salmonella’s major virulence factors were found in all isolates encoding partially chromosomal *Salmonella* pathogenicity island SPI-1 and SPI-2 including two Type 3 Secretion Systems (T3SS1-T3SS2) with exclusive and translocate effectors, and one Type 8 Secretion System (T8SS). Additionally, adherence, membrane, flagellar and chemotaxis kinase operons, and metal homeostasis pathways genes were described for all isolates. MGE related virulence genes gogB and grvA were observed in all STCs 2 and 3, and one isolate from STC 1 (Table S2).

All results of *Salmonella* Typhimurium could be explored at the microreact project (https://microreact.org/project/ty7n1isZqUKyu4ehFUvx1W-salmonellaentericatyphimurium-from-the-national-surveillance-in-colombia-as-one-health-approach).

### Salmonella Heidelberg

#### Relatedness analysis

*Salmonella* Heidelberg isolates from food (n=20) showed ten unique PPs with DSI values above 0.69, three most frequent PPs in food interface JF6.X01.0010, JF6.X01.0019 and JF6.X01.0027 (13/20) showed DSI values above 0.87 (figure 3a). Food interface PP JF6.X01.0010 (5/20) was clonally related with a clinical interface PP JF6.X01.0001 (1/3) (DSI=1.0) and close related, with a band of difference, from PP JF6.X01.0002 (1/3) (DSI=0.97) in the PulseNet National database at INS (figure 3b). *S*. Heidelberg Cluster -1 (ICA JF6.X01.0010 - INS JF6.X01.0001 - JF6.X01.0002) was defined. *S*. Heidelberg belonging to Cluster -1 were recovered in Bogotá D.C (5/7), Santander (1/7) and Boyacá (1/7) (Table S1). Other food isolates non-related to clinical interface were recovered from Bogotá (9/20), Tolima (2/20), Valle del Cauca (1/20), Córdoba (1/20), Cesar (1/20) and Santander (1/20) (S. Table 3). Four epidemiological variables from 20 food and 3 clinical *S*. Heidelberg isolates were compared, defining PSC in 0.73 (p<0.000). Comparison determined clonal relation in 4 food and 1 clinical isolates that exhibited AMR profiles Te, CipTe, CipLvxTe, FoxCipTe and NaTe from Bogota during 2010-2011 (Table S1).

Genomic comparison based on variants calling in *S*. Heidelberg was generated mapping to reference (NC_011083.1) 20 genomes: food (n=9) and clinical (n=11) isolates. A total of 11 isolates (58%) of *Salmonella* Heidelberg food-human interfaces were included based on SNPs distance (∼60 SNPs) resulted from the serovar phylogeny analysis: food (n=9) and clinical (n=2) isolates (figure 3c). All sequence types described for this serovar belong to CG 15 including ST15 (10/11) and ST7412 (1/11). A single *Salmonella* Heidelberg Clade (SHC1) was described, similarly to PFGE cluster -1 (ICA JF6.X01.0010 - INS JF6.X01.0001 - JF6.X01.0002), mentioned above. Isolates belong to SHC1 were recovered between 2009 - 2017 in Bogotá (7/11), Santander (2/11), Antioquia (1/11) and Norte de Santander (1/11). Two related genetic events were observed in three departments in 2009 and 2017, differentiated by 19 SNPs and 33 SNPs of distance between food and clinical interfaces, respectively (figure 3c; Table S2).

#### Antimicrobial resistance

. All food isolates showed resistance to at least one antibiotic family: b-lactam, b-lactamase inhibitor, fluoroquinolone, folate pathway inhibitor, tetracycline, aminoglycosides and nitrofurantoin. All five isolates belong to Cluster -1 from Bogota and Boyacá showed resistance to at least one antibiotic family: ß-lactam, ß-lactamase inhibitor, fluoroquinolone and tetracycline, with AMR profiles: Te (1/5), CipTe (1/5), CipLvxTe (1/5), FoxCipTe (1/5) and NaTe (1/5) (Table S1). MDR profiles were observed in two food isolates non-related to clinical interface from Santander and Bogotá: AmpCtxNaTe and AmcAmpCzCtxFoxCroTzpCipTe. Antimicrobial resistance determinants genes to aminoglycosides, ß-lactams, trimethoprim, fosfomycin, ammonium quaternary, quinolone, streptothricin, sulphonamides and tetracycline, and some mutations associated with quinolone resistance gyrA(S83F) and parC(T57S) were observed in SHC1 isolates. Incompatibility groups IncI and IncX were found in all SHC1 isolates and the other eleven plasmid incompatibility groups were described less frequently (Table S2).

#### Virulence factors

Major virulence factors for *Salmonella* were found in almost all SHC1 isolates encoding partially chromosomal *Salmonella* pathogenicity island SPI-1 and SPI-2 including two Type 3 Secretion Systems (TTSS1-TTSS2) with exclusive and translocate multiple effectors, and one Type 8 Secretion System (T8SS). Additionally, adherence, membrane, flagellar and chemotaxis kinase operons, and metal homeostasis pathways genes were described for all isolates (Table 2; figure 3c). Yersinia High Pathogenicity Island (HPI) was detected in all isolates belonging to SHC1, including ybt (SXQPA)(UTE), irp1, irp2 and fyuA. Additionally, grvA, gogB and cdtB genes were observed occasionally in some isolates from SHC1 (Table S2).

All results of *Salmonella* Heidelberg could be explored at the microreact project (https://microreact.org/project/rwdTW9g5esRp3V6wkodz7w-salmonella-enterica-heidelberg-from-the-national-surveillance-in-colombia-as-one-health-approach).

### Salmonella Paratyphi B dT+

#### Relatedness analysis

*Salmonella* Paratyphi B dT+ (n=23) from food showed thirteen unique PPs with DSI values from 0.53, the most frequent PPs were JKX.X01.0009 (3/23), JKX.X01.0013 (4/23) and JKX.X01.0014 (5/23). Comparison of PPs from human and food interfaces did not reveal any clonal or close relation. *S*. Paratyphi B dT+ isolates were recovered from Bogotá D.C (17/23), Atlántico (3/23), Caldas (2/23) and Tolima (1/23).

#### Antimicrobial resistance

All *S*. Paratyphi B dT+ isolates recovered from food were MDR to several antibiotic families: quinolones, tetracycline, ß-lactam, ß-lactamase inhibitor, fluoroquinolone, aminoglycoside, folate pathway inhibitor and nitrofurantoin. Two MDR profiles AmcAmpCzoCazCroCtxXnlFoxNaCipLvxEnrStrNitTeSxt and NaSxtTeStr were observed in Atlántico and Bogotá, and Valle del Cauca, respectively.

There was no similarity between *S*. Paratyphi B dT+ isolates from food and clinical interfaces between 2010-2011, therefore, this serovar was excluded from the genomic analysis (Data no showed).

## Discussion

The retrospective comparison of *Salmonella* spp. from the food and clinical interfaces as proposed in this study represents the best approximation to integrated genomic analysis of *Salmonella* foodborne in Colombia from the OH approach. This study adds evidence of the benefits of WGS over PFGE on *Salmonella* spp. surveillance and outbreak investigations across the different interfaces, following most certainly the establishment of relatedness in foodborne chains, especially in bacteria with clonal behaviour as *Salmonella* Enteritidis. Likewise, the early and precise detection of emerging foodborne risks as the spread of multi-drug resistance and virulence convergency in bacteria pathogens (44). Furthermore, for the first time in Colombia, we provided evidence of the relatedness of *Salmonella* food isolates obtained from poultry chain and *Salmonella* isolates which are causing invasive disease for a prolonged time and geographical space, especially in childhood and elders at the clinical interface.

Different molecular techniques have been applied to *Salmonella* surveillance and outbreak’s investigations in Colombia and in other world regions (29,45–47). WGS has become the gold standard procedure for foodborne surveillance in public health laboratories in High-Income Countries, showing huge advantages and the best resolution on investigations of pathogens (20,48,49). The COVID-19 pandemic provides a unique opportunity to sensibilize general audiences, decisions and policymakers in LMICs about the applicability of WGS on infection disease surveillance and monitoring (50). Recently, Europe, Asia, Africa and the Americas reported *S*. Enteritidis Complex Group 11 foodborne outbreaks associated with poultry meat products, confirming prevail global importance of this ancestral type in human infections. Application of WGS to *Salmonella* Enteritidis most prevalent serovar in Colombia, allowed us to understand the limitations of current molecular surveillance (PFGE), resolving close genetic relations across the interfaces in 8 clades, more than third as many clusters as PFGE, and with greater additional useful pathogenic data. The *S*. Enteritidis clades (SECs) associated with poultry in this study were more significant than in similar studies, suggesting higher complexity in contamination sources in Colombia (51–54). Therefore, having a powerful toolbox for the certain assessment of genetic relations across interfaces can help establishing dissemination routes and supporting real-time measurements and control actions in public health. Similar study in the PulseNet USA showed that WGS reduce 34% of the cases misplaced in outbreaks, revealing a higher percentage of confirmed outbreak cases by WGS clustering (78%) than PFGE clusters (46%)(55). Nevertheless, implementation of genomics in LMICs has multi-factor challenges, especially high cost, complexity, infrastructure and scarce trained human resource, which need to be kept in mind for the implementation of genomic surveillance domestically (40)(56). Other challenges of WGS are the interpretation and communication of results between different stakeholders, that is not straight-forward as PFGE typing method that have a stable nomenclature, and the less time on WGS outcome than the traditional PFGE as routine on laboratories (55)(57). Nonetheless, the bioinformatic versioned pipelines and the easy to share interactive WGS reports used in this study could help to overcome these challenges in the national reference laboratories in Colombia and elsewhere.

Moreover, three zoonotic high-risk clades identified in this study in *Salmonella* Typhimurium (STCs) exhibited clonal dissemination, similar to that obtained for PFGE, nevertheless, WGS exhibited the pooling of other patterns into the clades. Also, diverse virulence factors, AMR determinants to critical antibiotic families and complex mobile genetic platforms were observed, which confirms the relevance of this serovar as foodborne and in AMR dissemination in Colombia. *S*. Typhimurium outbreaks are frequently related to other types of animal food sources such as swine and cattle, more that poultry (58,59). Due the lower number of *S*. Typhimurium food isolates described in this study, the dissemination route could be related also to cross-contamination at retail stores with additional sources as other animal meats, humans, transport and equipment (59). WGS increased the discrimination power to differentiate the STCs, providing specific biomarkers that could be used for routine molecular tracing or rapid detection methods of these high-risk clades in LMICs at NMRL surveillance programs and monitoring (60). Additionally, this is the first report in food of mcr-5.1 gen in Colombia observed in STC-1 isolates that is associated with resistance to colistin, which is the last resource to control carbapenem resistance Gram-negative bacteria. Colombia is a Carbapenem resistance endemic country in other critical pathogens such as *Klebsiella pneumoniae, Acinetobacter baumanii* and *Pseudomona aeruginosa*, therefore colistin has been gaining prominence on antimicrobial control and surveillance (31,61,62).

WGS also allowed us to observe in the *Salmonella* Heidelberg clade, similar to PFGE discrimination since 2010, the convergence of complex AMR profiles and virulence factors as yersiniabactin located in *Yersinia* High Pathogenicity Island (HPI). In 2021, HPI was described for the first time in *Salmonella* serovar from Heidelberg ST-15 in Brazil. Previously, HPI was described in other *Salmonella* serovars in clinical and turkey production interfaces by Germany and USA. Similar reports in other species such as *E*. coli have been related HPI in the increasing virulence and resilience in the host, besides increasing pathogenicity in poultry production systems (63–65). In Colombia, *S*. Heidelberg has been reported as the second most prevalent serovar in poultry production, but it is less common in human invasive infections (28,29). Nevertheless, this serovar should be monitored in the national surveillance due to the observed pathogenic potential and the relevance as an invasive bacteria in other countries (24,66). Likewise, *S*. Paratyphi B dT+ (Java) has been reported as the most prevalent serovar recovered in Colombia at poultry farms, slaughterhouses and retail stores, but it was scarce in the clinical isolates included in this study (67). Nonetheless, it should be monitoring as a public health concern in Colombia, due to the high performance as multi-drug resistance reservoir and the increasing of paratyphoid infectious reports in the clinical (24).

Finally, although *Salmonella* from the clinical was defined with the highest complexity of AMR profiles, suggesting higher selective pressures at clinical and community environments, and *Salmonella* from poultry origin was found the primary carrier of acquired virulence factors (68–70). The use of WGS allowed us to expose the dissemination in food of pathogenic bacteria as *Salmonella* and the drug-resistance and virulence, and their close relation with human invasive disease for 20 years in Colombia. Other food sources could be involved in human invasive Salmonellosis infections in Colombia, however, this study provided evidence of foodborne clades associated with the poultry chain. Nevertheless, it is essential to provide regular WGS data from the poultry and other food chains but from other interfaces to generate a much more comprehensive analysis of *Salmonella* dissemination and its zoonotic dynamics in Colombia (71)(72)(73). WGS makes feasible the integration of data from foodborne clades domestically and globally, allowing institutions to respond more efficiently to health threats as they arise. Also, increases the opportunities for actionable effective strategies in regulatory institutions to contain infectious diseases and reducing the risks in food to the population.

### Biological material

Food strains are preserved in the *Salmonella* collection of the Colombian Integrated Program of Antimicrobial Resistance Surveillance COIPARS-AGROSAVIA deposited in custody at National Veterinary Diagnostic Laboratory (LNDV) in National Agriculture Institute (ICA); The clinical strains are preserved at National Microbiology Reference Laboratory at National Health Institute (INS).

### Bias

This study had a possible bias, related to 1) The differences between the time range in WGS analysis, where food isolates included were recovered from two years (2010-2011), and clinical isolates recovered for a longer term of ten years (1997-2017), 2) Over-representation of clinical isolates compared with food isolates (∼10%), could mask new un-reveal risks of foodborne *Salmonella*. 3) Clinical isolates selection for WGS was not based on the initial molecular comparison using PFGE.

*S*. Heidelberg and *S*. Paratyphi B dT+ isolates included in this study represent a sample from isolates collected in food at retail stores in our previously study, this convenience selection was based on available data from clinical isolates, where historically *Salmonella* Heidelberg and Paratyphi B dT+ prevalence was extremely low (4 samples) focusing the initial study on public health concern *Salmonella* serovars in Colombia.

## Data Availability

Availability of sequence data. The raw sequencing data generated from this study were deposited at the EMBL European Nucleotide Archive (ENA) repository under the project accession numbers PRJEB35182 and PRJEB47910.

https://www.protocols.io/view/ghru-genomic-surveillance-of-antimicrobial-resista-bpn6mmhe

https://www.ebi.ac.uk/ena/browser/view/PRJEB35182

https://www.ebi.ac.uk/ena/browser/view/PRJEB47910

## Abbreviation

(Amp): Ampicillin
(C): Chloramphenicol
(Amc): Amoxicillin-clavulanic acid
(Te): Tetracycline
(Str): Streptomycin
(Cip): Ciprofloxacin
(Lvx): Levofloxacin
(Na): Nalidixic acid
(Nit): Nitrofurantoin
(Czo): Ceftazidime
(Ctx): Cefotaxime
(Cro): Ceftriaxone
(Fox): Cefoxitin
(Tzp): Tazobactam-piperacillin
(Col): Colistin
(ICA): Instituto Colombiano Agropecuario.

## Acknowledgments

Dr. McAllister Tafur from Andean community for his advice and funding support in the early stages of the project at ICA laboratory. Dr. Enrique Perez from the Pan-American Health Organization (PAHO) for his advising, support and global networking, Dr. Walid Alali from the Center of Food Safety, University of Georgia – USA for his technical and financial support in the earlier studies of retail stores, and Dr. Isabel Chinen from the Instituto Malbrán-Argentina, regional head from the international foodborne network PulseNet LA&C .for technical support and regional cooperation.

---

**Table S1.** Similarity analysis of *Salmonella* spp. isolates from food and clinical compared by Serovar, PFGE profile, Location and Antimicrobial resistance (AMR).

| Serovar | # Total isolates | PFGE_food_profile | PFGE_clinic_profile | # Total isolates by serovar /PFGE | Location | # Total isolates by serovar/PFGE /location | AMR | # Total isolates by serovar/ PFGE/location/ AMR | # Related isolates food | # Related isolates clinical | Pearson Similarity coefficient (p<0.000) |
| --- | --- | --- | --- | --- | --- | --- | --- | --- | --- | --- | --- |
| S. Enteritidis | 324 | X01.0001 | X01.0001 | 211 | Bogota (n=75)<br>Atlantico (n=3)<br>Arauca (n=1)<br>Others (n=132) | 75 | None /AmpTeNaCip | 61 | 23 | 38 | 0.86 |
|  |  | X01.0006 | X01.0038 | 61 | Bogota (n=29)<br>Valle del Cauca (n=6).<br>Risaralda (n=2)<br>Arauca (n=1)<br>Others (n=23) | 37 | None | 33 | 18 | 15 |  |
|  |  | X01.0008 | X01.0068 | 2 | Bogota (n=2) | 2 | None | 2 | 1 | 1 |  |
| S. Typhimurium | 430 | X01.0007 | X01.0197 | 4 | Bogota (n=4) | 4 | CAmcAmpTeStr | 4 | 1 | 3 | 0.89 |
|  |  | X01.0004 | X01.0237 | 12 | Antioquia (n=6)<br>Bogota (n=3)<br>Boyaca (n=3) | 3 | TeStr | 2 | 1 | 1 |  |
| S. Heidelberg | 23 | X01.0010 | X01.0001/2 | 7 | Bogota (n=5)<br>Santander (n=1)<br>Boyaca (n=1) | 6 | CipTe | 5 | 4 | 1 | 0.79 |

\*Antibiotics abbreviation: Ampicillin (Amp); Chloramphenicol (C); Amoxicillin-clavulanic acid (Amc); Tetracycline (Te); Streptomycin (Str); Ciprofloxacin (Cip)

**Table S2.**
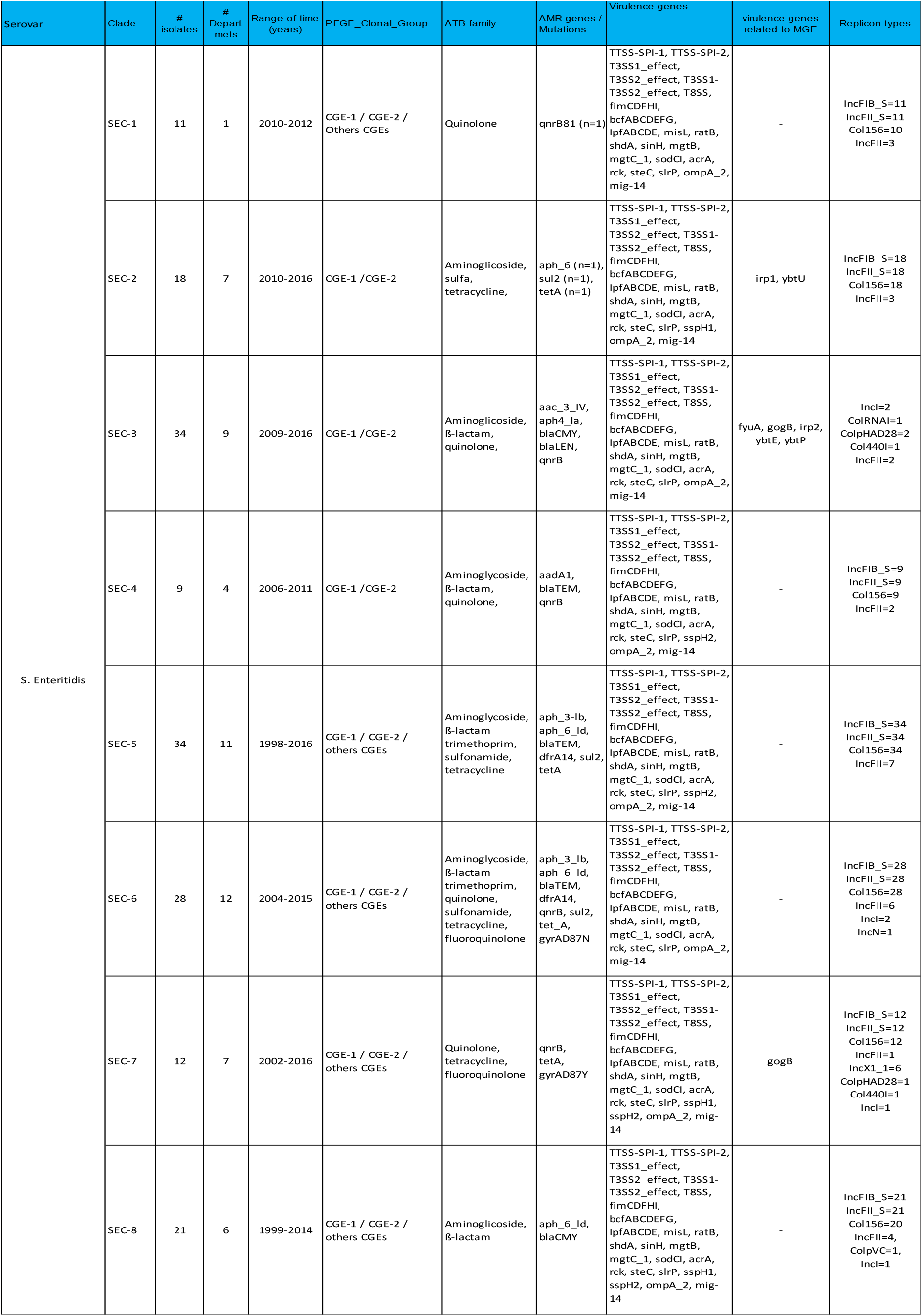
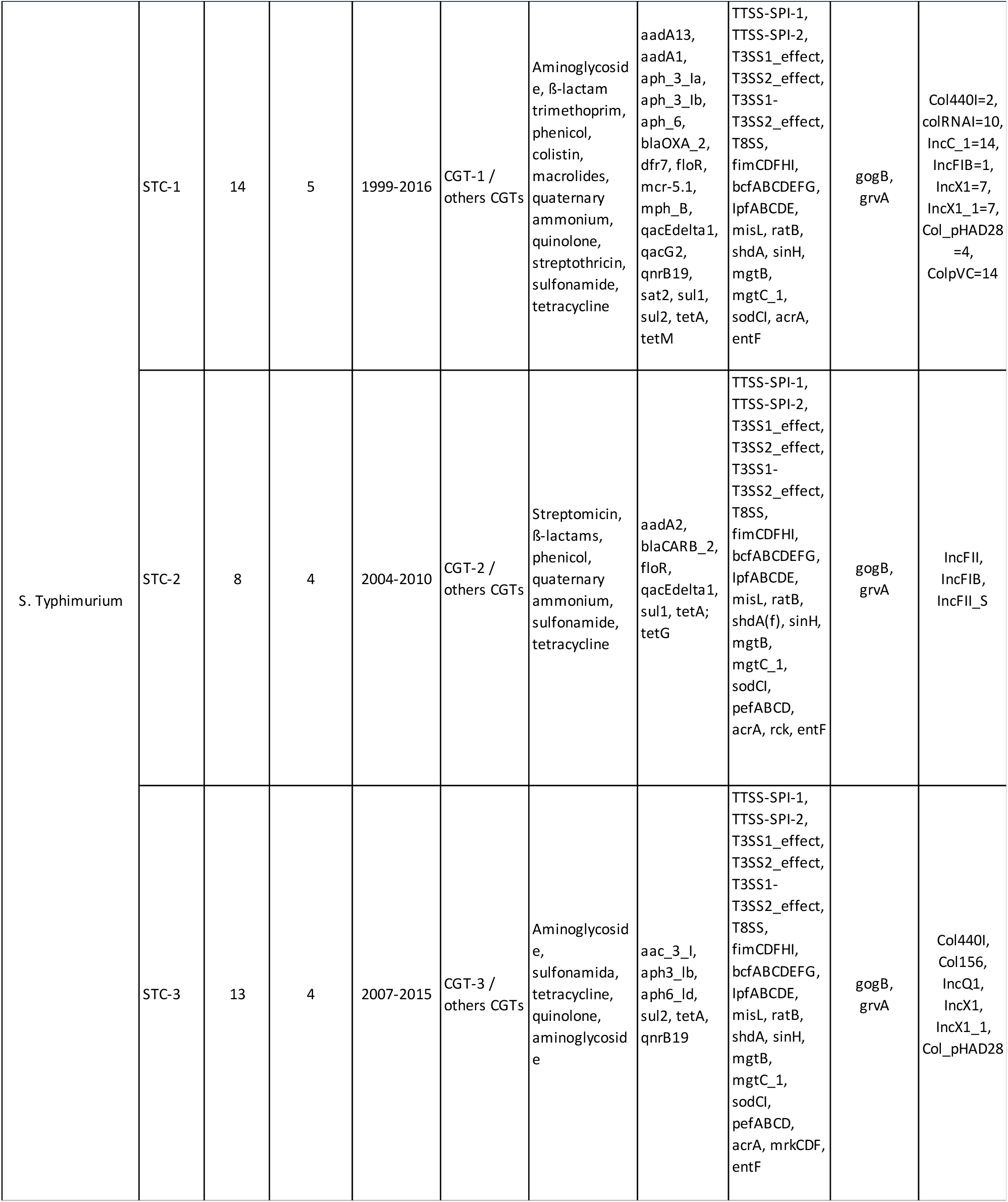
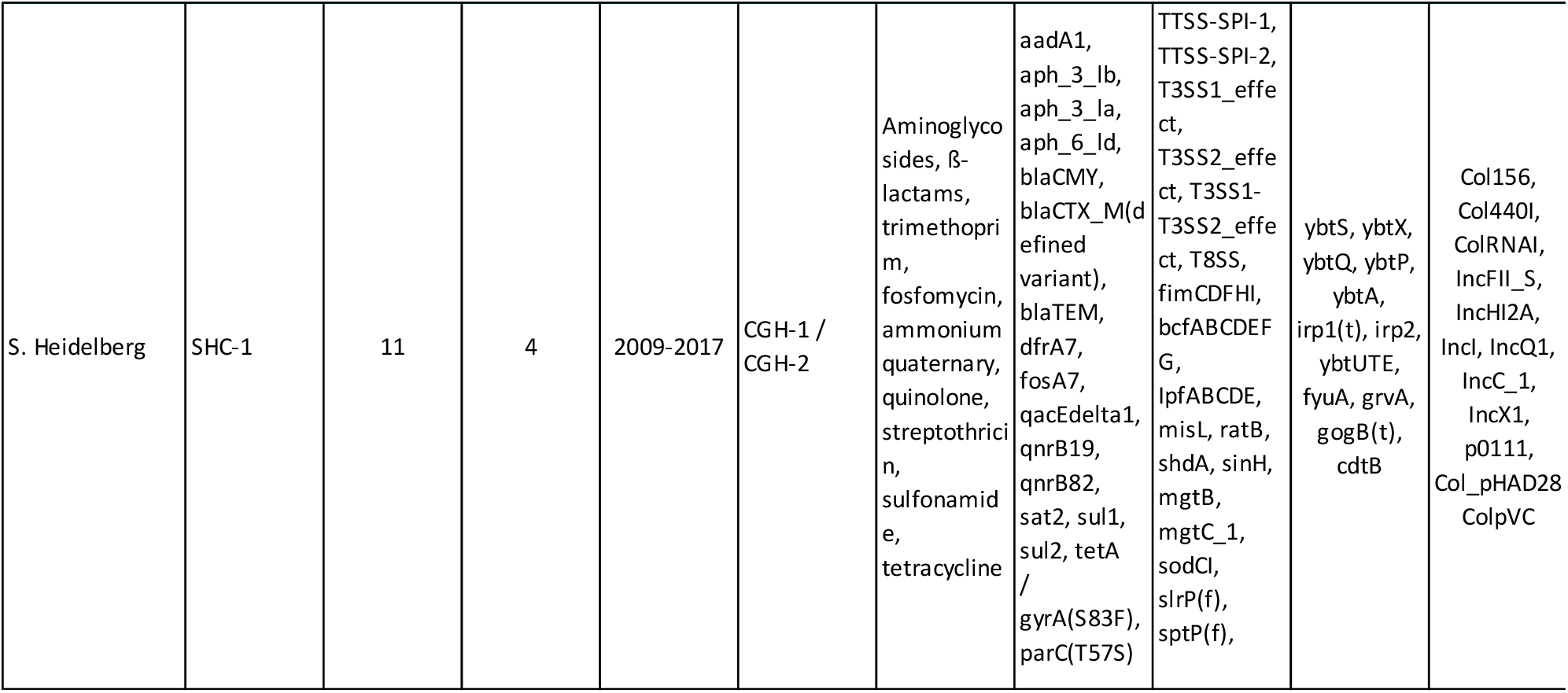
Summary of WGS results by serovar, clades, antimicrobial resistance, virulence and MGE.

**Figure 1a.**
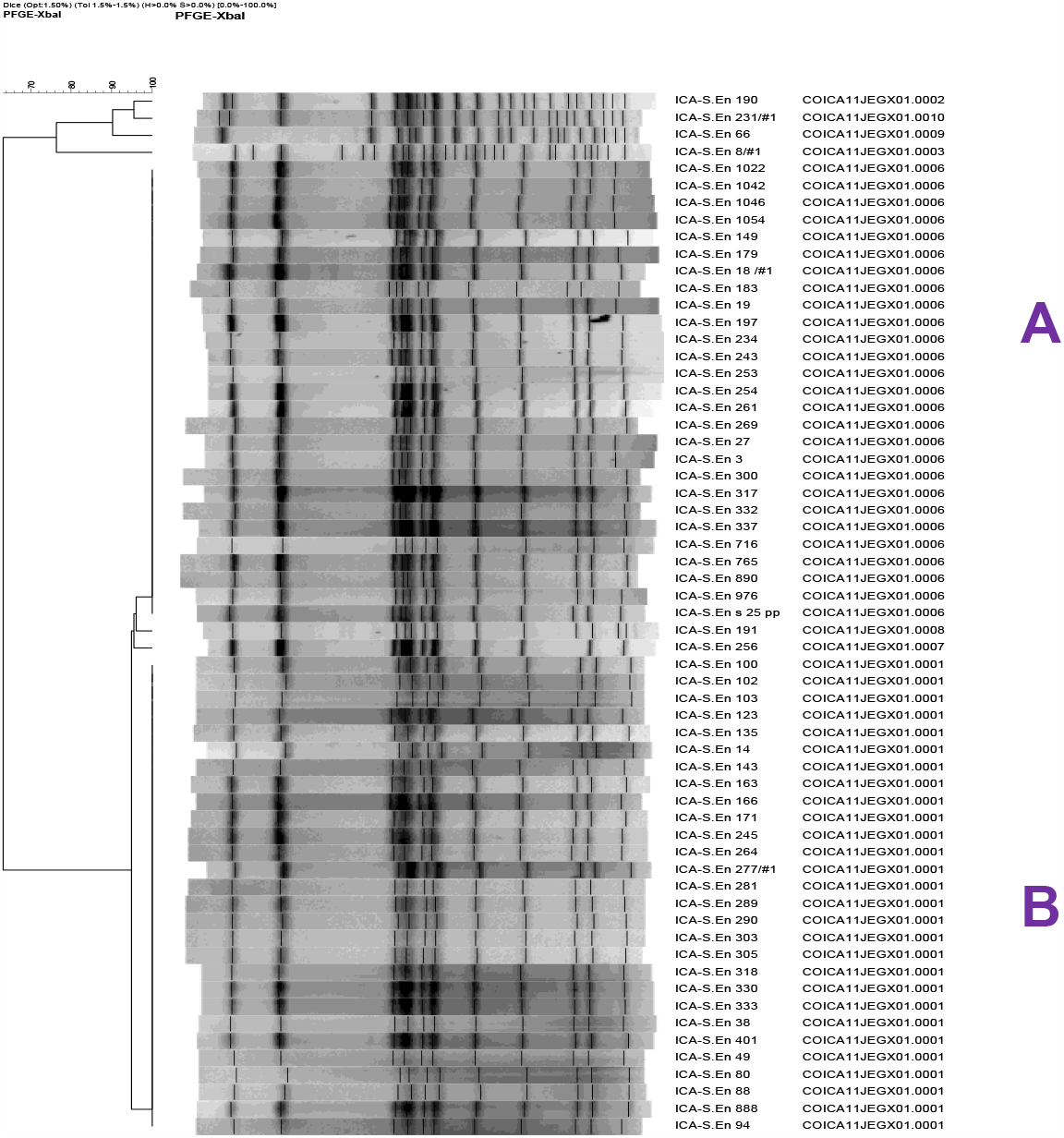
Similarity clustering of *Salmonella* Enteritidis PFGE patterns from food samples using DICE similarity index and clustering by UPGMA. Two clonal clusters were identified in food isolates: A) Isolates with pattern ICA JEG.X01.0006 and B) Isolates with pattern ICA JEG.X01.0001

**Figure 1b.**
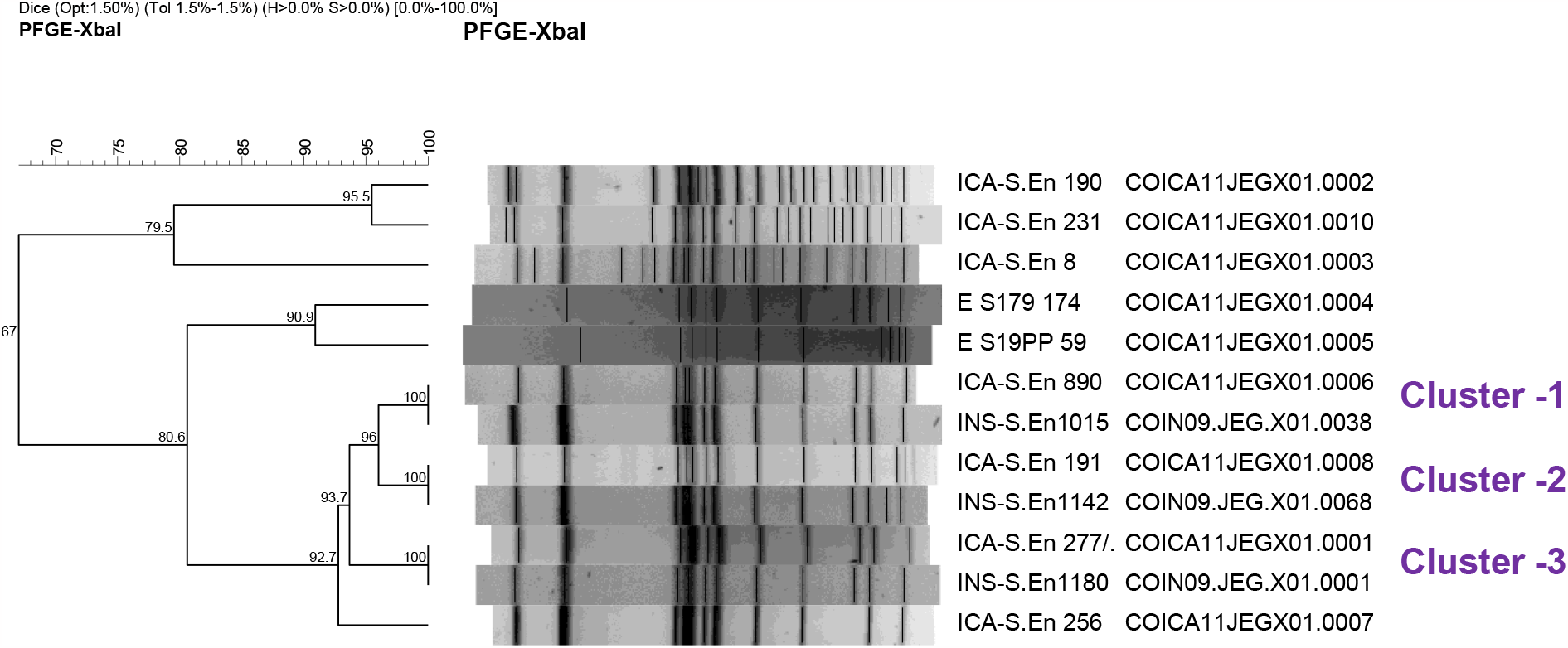
Comparison of related unique *Salmonella* Enteritidis PFGE patterns from clinical (INS) and food (ICA), using DICE similarity index and clustering by UPGMA. Three pairs of patterns were found clonal (SI%:100) related: Cluster -1 JEG.X01.0006 - JEG.X01.0038, Cluster -2 JEG.X01.0008 - JEG.X01.0068 (singleton) and Cluster -3 JEG.X01.0001 - JEG.X01.0001.

**Figure 1c.**
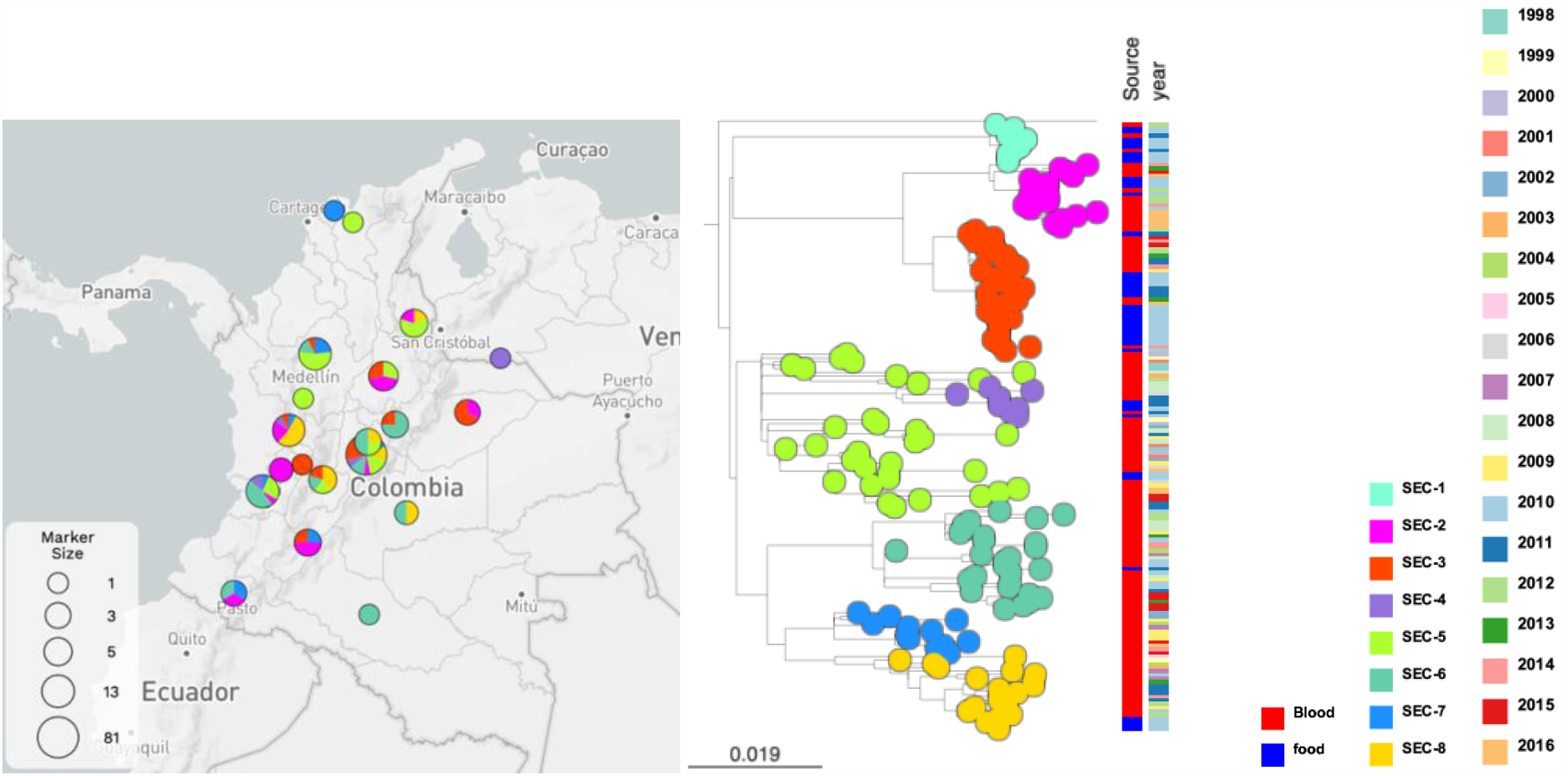
Geographic distribution and phylogeny inference by clade of *Salmonella* Enteritidis (https://microreact.org/project/hjw5geMmKWAAgKxhA9Y3QX-salmonellaentericaenteritidis-from-the-national-surveillance-in-colombia-as-one-health-approach).

**Figure 1d.**
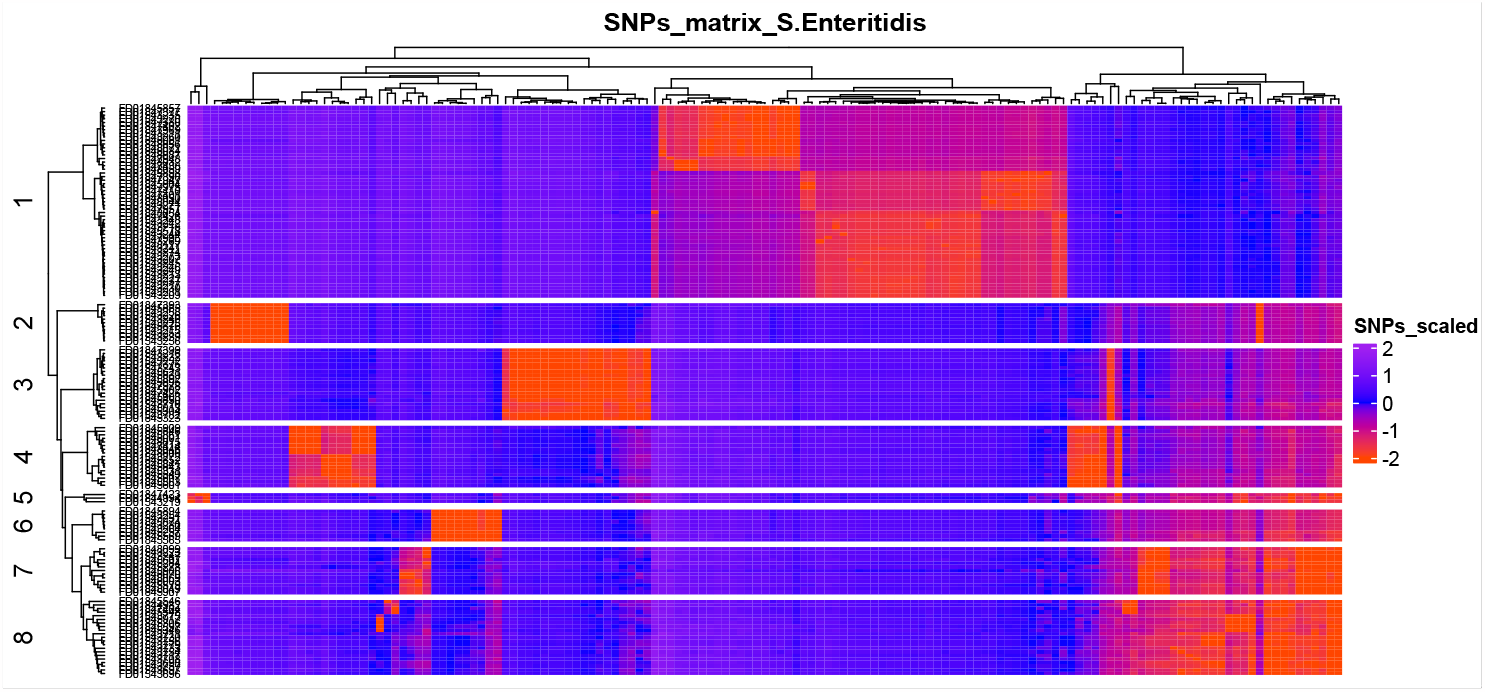
SNPs matrix distribution of *Salmonella* Enteritidis clades

**Figure 2a.**
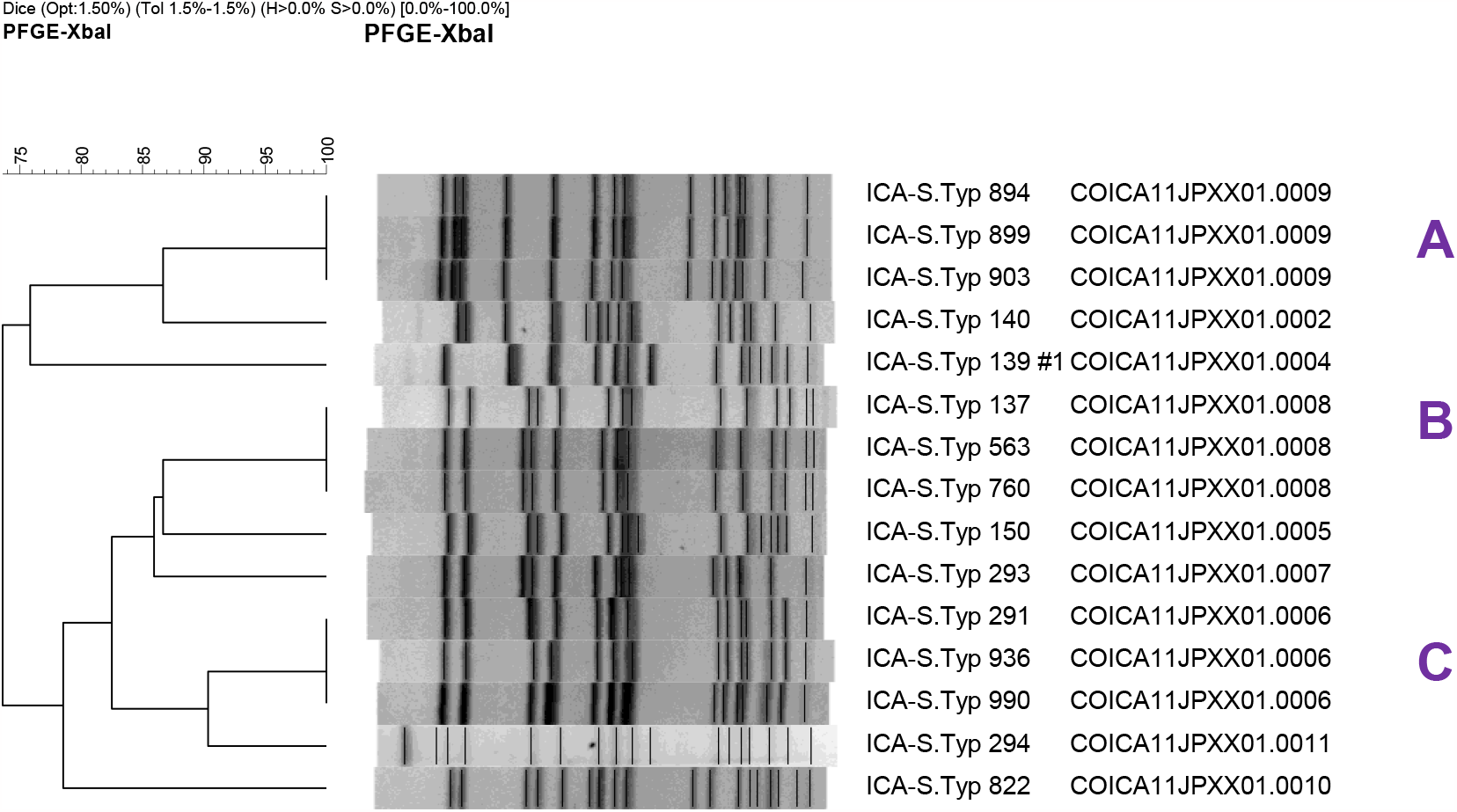
Similarity clustering of *Salmonella* Typhimurium PFGE patterns from food samples using DICE similarity index and clustering by UPGMA. Three clonal clusters were identified in food isolates: A) Isolates with pattern ICA11.JPX.X01.0009, B) Isolates with pattern ICA11.JPX.X01.0008 and C) Isolates with pattern ICA11.JPX.X01.0006.

**Figure 2b.**
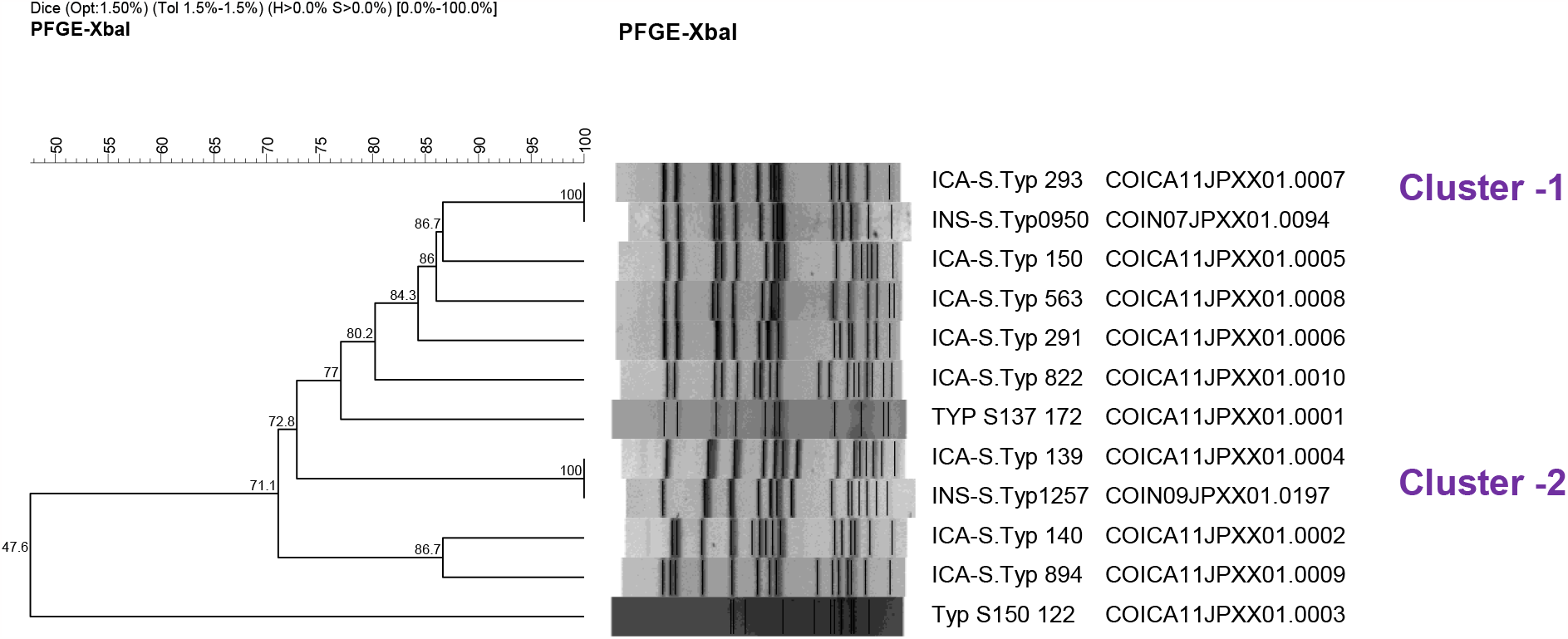
Comparison of unique *Salmonella* Typhimurium PFGE patterns from clinical (INS) and food (ICA) using DICE similarity index and clustering by UPGMA. Two pairs of patterns were found clonal (SI%:100) related: cluster -1 JPX.X01.0007 - JPX.X01.0094 and cluster -2 JPX.X01.0004 - JPX.X01.0197.

**Figure 2c.**
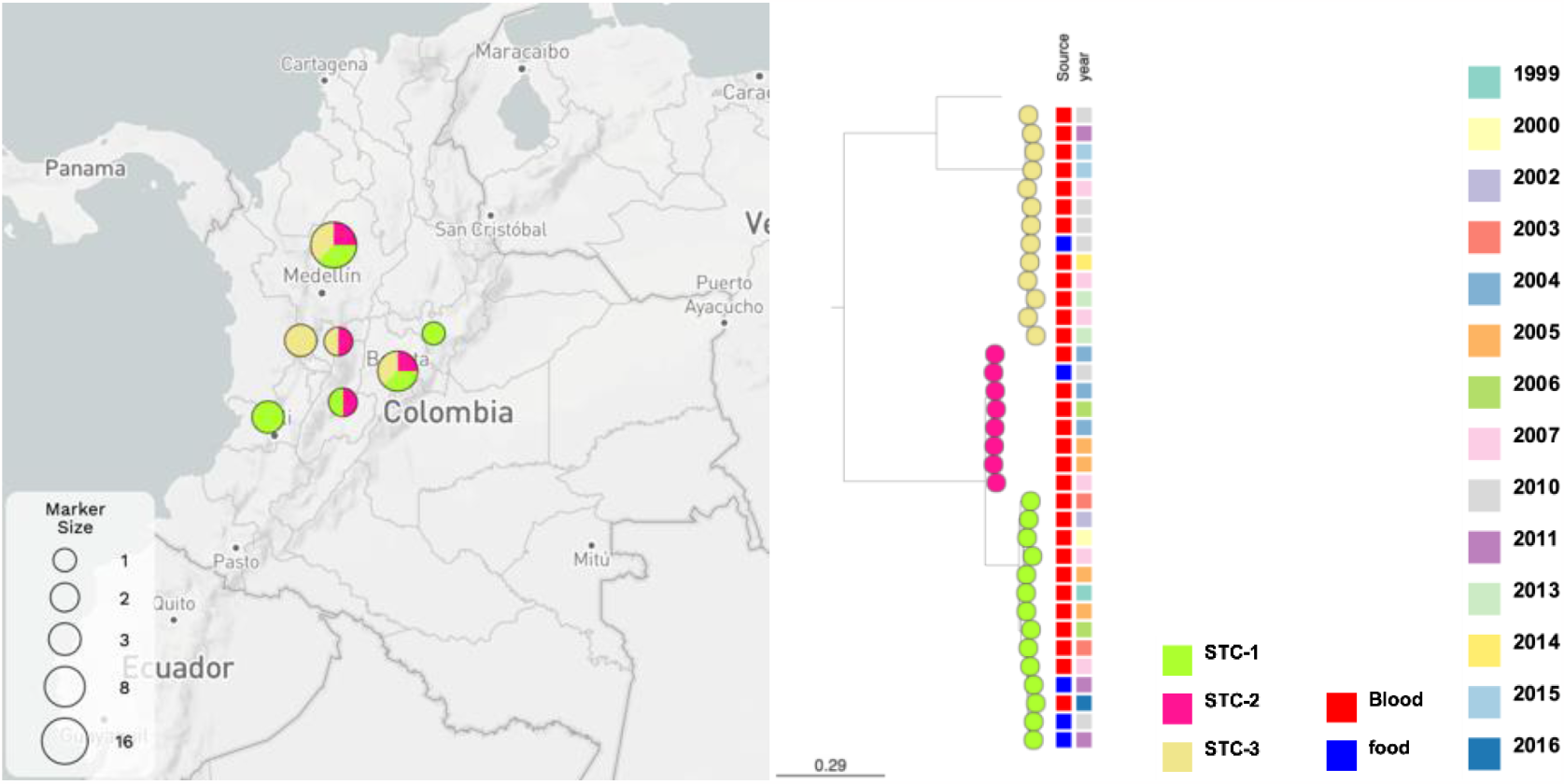
Geographic distribution and phylogeny inference by clade of *Salmonella* Typhimurium samples (https://microreact.org/project/ty7n1isZqUKyu4ehFUvx1W-salmonellaentericatyphimurium-from-the-national-surveillance-in-colombia-as-one-health-approach)

**Figure 2d.**
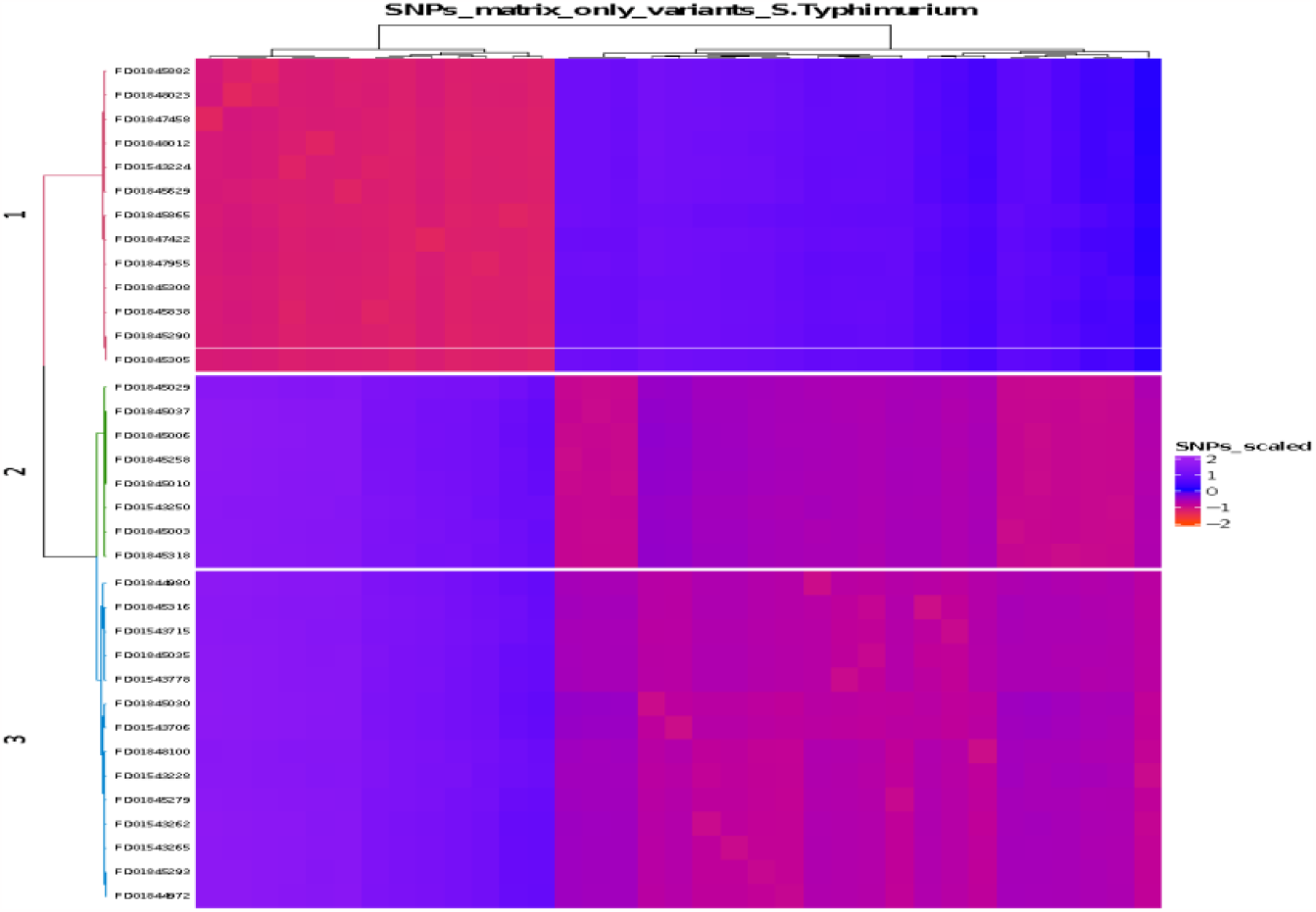
Visualization of SNPs matrix distribution of *Salmonella* Typhimurium clades

**Figure 3a.**
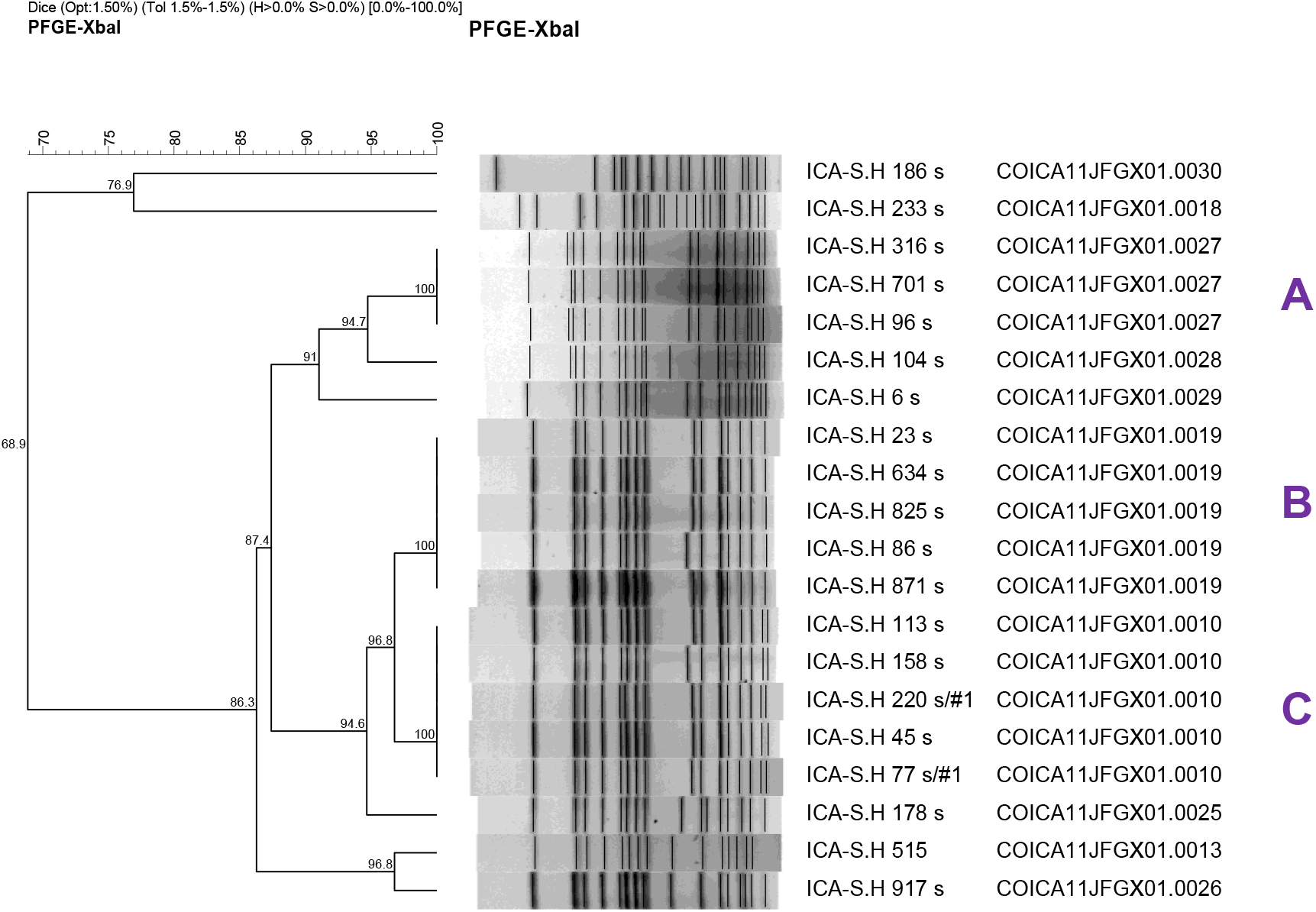
Similarity clustering of *Salmonella* Heidelberg PFGE patterns from food samples using DICE similarity index and clustering by UPGMA. Three clonal clusters were identified in food isolates: A) Isolates with pattern ICA JFG.X01.0027, B) Isolates with pattern ICA JFG.X01.0019 and C) Isolates with pattern ICA JFG.X01.0010.

**Figure 3b.**
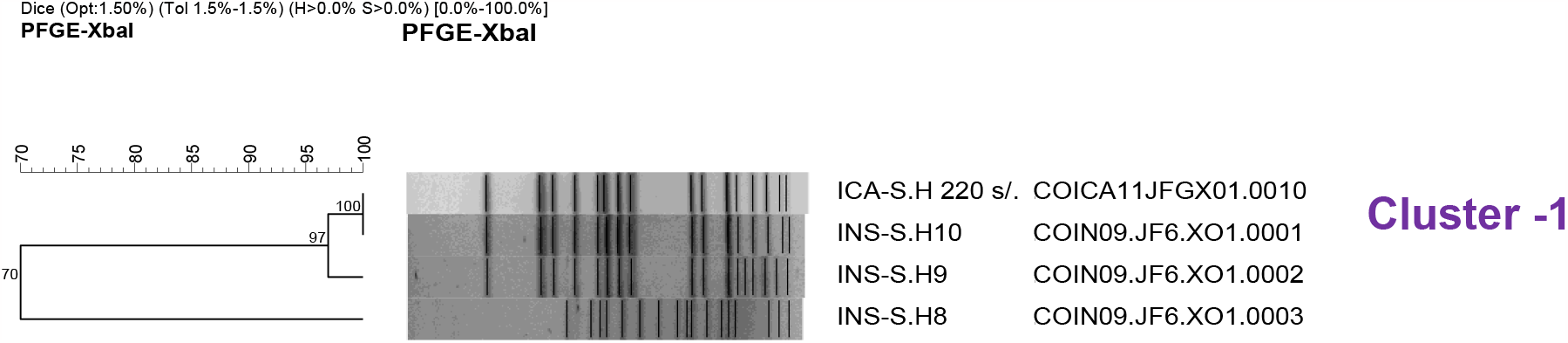
Comparison of unique *Salmonella* Heidelberg PFGE patterns from clinical and food using DICE similarity index and clustering by UPGMA. One pair of patterns was found clonal (DSI%=100) related: Cluster -1 INS JF6.X01.0001 – ICA JFG.X01.0010. Another clinical pattern was found close related (DSI%=97) to the clonal cluster: JF6.X01.0002

**Figure 3c.**
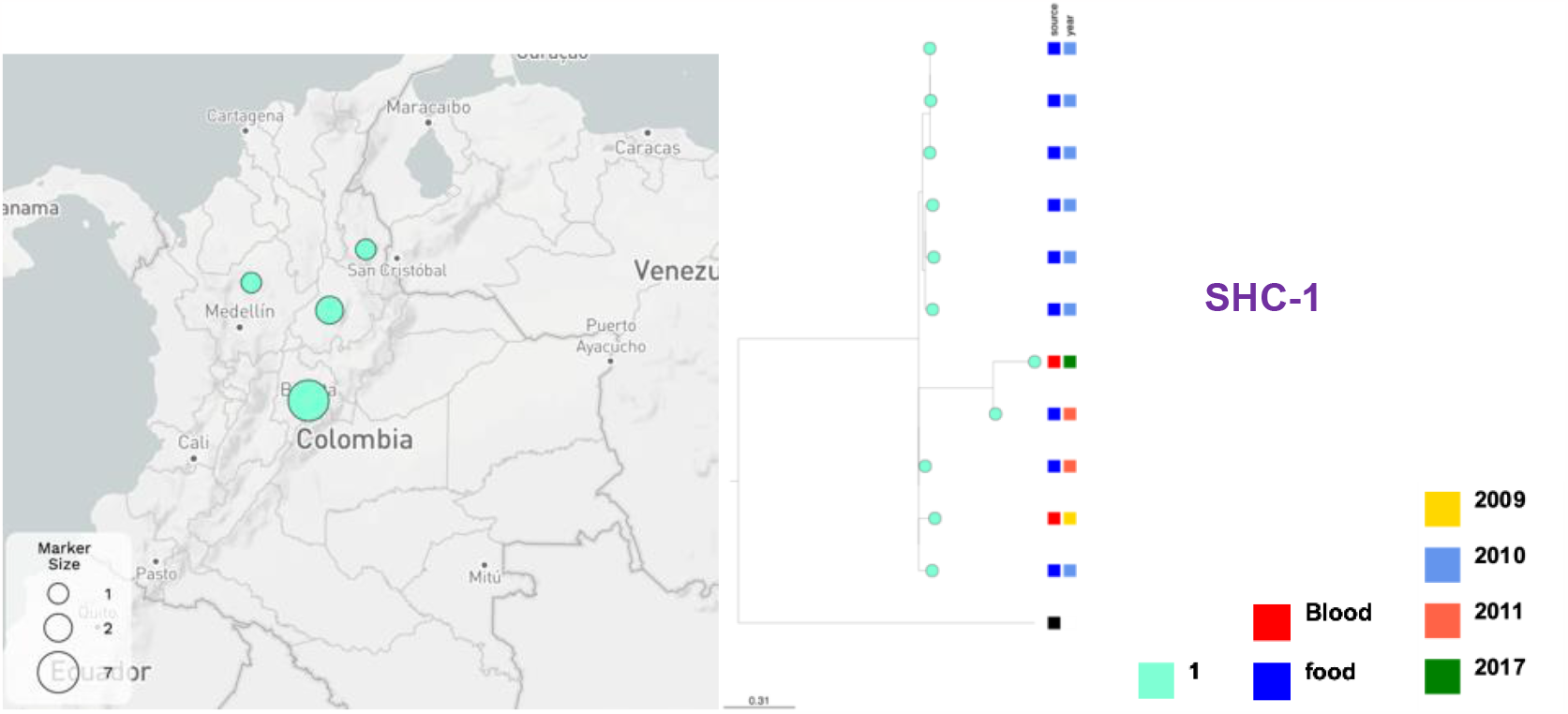
Geographic distribution and phylogeny inference by clade of *Salmonella* Heidelberg (https://microreact.org/project/rwdTW9g5esRp3V6wkodz7w-salmonella-enterica-heidelberg-from-the-national-surveillance-in-colombia-as-one-health-approach).

**Figure 3d.**
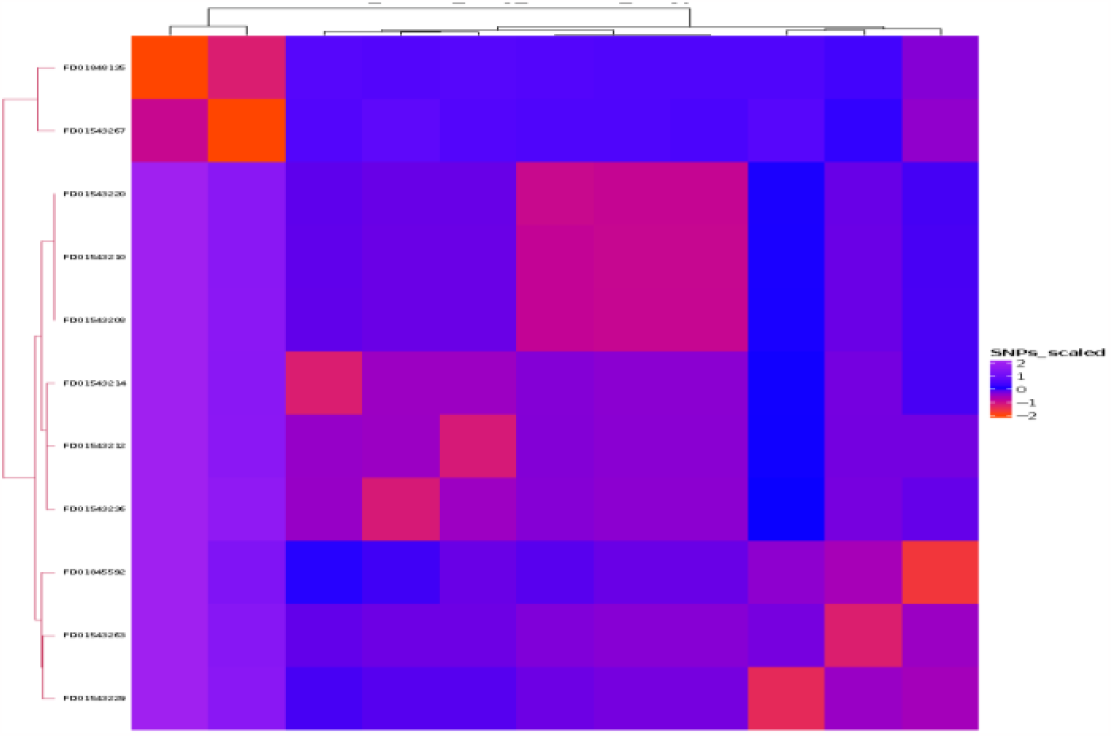
Visualization of SNPs matrix distribution of *Salmonella* Heidelberg clade

